# Increase of *Clostridioides difficile* PCR ribotype 002 infection coincides with decreased disease severity in hospitalized patients in the Netherlands

**DOI:** 10.1101/2024.11.11.24316883

**Authors:** A Baktash, C Harmanus, J Goeman, W Brennan, M Cormican, J Corver, EJ Kuijper, WK Smits

## Abstract

*Clostridioides difficile* PCR ribotype (RT) 002 is the second most prevalent RT in the Netherlands. In 2019, an increase in *C. difficile* infections (CDI) due to a clonal RT002 strains was reported in Ireland. In retrospect, a significant increase in the proportion of CDI due to RT002 was detected in the Netherlands between May 2017-May 2018 (11.9% of 683 cases vs 6.5% of 865 in the previous year). Our aim was to analyze and characterize CDI due to RT002 over time and to assess whether RT002 isolates in the Netherlands were related to the Irish RT002 isolates.

Over the period May 2009-April 2021, Dutch patients with CDI due to RT002 showed less severe CDI (22%) in comparison with hypervirulent RTs (RT027; 29% and RT078; 25%), similar to grouped data from other non-hypervirulent ribotypes (21%). However, RT002-associated CDI prior to the increased proportion (2009-2017) showed CDI severity similar to CDI associated with hypervirulent types, whereas this was no longer the case after the increase (2017-2021). Using core genome MLST, Dutch isolates were found not to be related to the Irish outbreak strain. Several Dutch strains clustered together without an evident epidemiological link, suggesting recent emergence.

**Importance:** *Clostridioides difficile* is a significant cause of hospital-associated infections, making it relevant to understand the emergence and spread of different ribotypes. This study highlights a rise in cases of PCR ribotype 002 (RT002) in the Netherlands, suggesting changes in its prevalence. Despite the increase in RT002 infections, the severity of disease appears to have decreased and is lower compared to more virulent ribotypes. Further research is needed to understand the factors driving this change. Additionally, this study underscores some limitations of core genome multi-locus sequence typing (cgMLST). cgMLST is valuable for identifying genetic relationships, however it cannot always distinguish between widespread recently emerged strains and local transmission chains, making it challenging to determine the exact sources of infection. Continued surveillance and genetic characterization of *C. difficile* are essential to better understand these trends and improve control measures.

## Introduction

*Clostridioides difficile* is an anaerobic, spore-forming, Gram-positive bacterium that causes infections varying from antibiotic-associated diarrhea to life-threatening pseudomembranous colitis (1). Hypervirulent PCR ribotypes such as RT027 and RT078 are associated with severe and recurrent CDI, but CDI and outbreaks are not limited to these ribotypes (1).

RT002 is one of the most common ribotypes in the Netherlands and in Europe (2). Besides one case report about *C. difficile* bacteremia due to RT002 in a patient from Belgium (3), little information is available on RT002-associated CDI in Europe, with most publications stemming from East-Asia where RT002 is highly prevalent. *In vitro* assessment of bacteriological characteristics of RT002 strains showed a higher amount of toxin A and toxin B production, sporulation and germination compared to strains belonging to other ribotypes (e.g. RT012, RT014 and RT046) (4). Comparable results were reported in another study from Hong Kong (5). RT002 also caused more weight loss and histological damage in a murine model of CDI (4). RT002 has been reported to be highly prevalent. For instance, RT002 is highly prevalent among nursing home residents in Hong Kong (5), (6). RT002 infections were associated with high morbidity and mortality in elderly patients (7). Patients with CDI due to RT002 showed an association with more usage of β-lactam antibiotics in the previous three months compared with controls (5). Furthermore, there was also an association with fluoroquinolone resistance (7).

Motivated by these reports, we assessed the prevalence of CDI due to RT002 in the Netherlands, and retrospectively noted an increase in the proportion of CDI due to RT002 between May 2017-April 2018, compared to previous years (11.9% vs 5.4-7.2% in the previous years).

In this study, we compare the clinical characteristics of CDI due to RT002 with those of CDI caused by both hypervirulent and non-hypervirulent ribotypes. Considering an increase in the proportion of CDI due to RT002 in the Netherlands in 2017, we also performed a sub-group analysis in which the strains were grouped based on isolation date. Finally, we assessed whether Dutch RT002 isolates were related to Irish clonal strains.

## Materials & Methods

### Clinical data collection

Clinical data collected from May 2009 until April 2021 (N=8080 strains PCR ribotyped) from the Dutch national CDI sentinel surveillance were used to analyze the clinical characteristics of CDI episodes due to RT002. All hospitalized patients (>2 years old) in participating hospitals (N=24 acute care hospitals) with clinical signs of CDI in combination with a positive test for *C. difficile* toxins or toxigenic *C. difficile*, were registered. The local laboratory chose the indication for testing for CDI, the assay and the algorithm that was used for diagnosis of CDI. CDI was described as severe if at least one of the following conditions was present; fever (≥ 38⁰C) and leukocytosis (>15*10^9^/L), dehydration and/or hypoalbuminemia (<20 g/L), bloody diarrhea and pseudomembranous colitis (8). A complicated CDI course was defined as: admission to the intensive care unit, the need for a surgical procedure and/or mortality within 30 days after diagnosis of CDI (either due to CDI or non-CDI-related) (8).

Clinical characteristics of CDI episodes due to RT002 were compared with the results of other RT groups. These groups were RT027 and RT078/126 (which are considered as hypervirulent strains), RT001 and RT014/020/295 (which are common non-hypervirulent strains in the Netherlands) and the other group (excluding the 4 previous strains). Ribotypes RT014/RT020/RT295 and RT078/RT126 that were difficult to distinguish from each other with PCR ribotyping, but not with CE-PCR ribotyping were combined in one group, since the data collecting period encompassed both methods. The data set was also split in two based on isolation date.

### Ethics

This observational study used data from the Dutch national CDI sentinel surveillance program, which has been in operation since 2009. The program collects microbiological and clinical data from all hospitalized patients diagnosed with CDI at participating hospitals across the Netherlands and was developed by the National Institute for Public Health. No additional data, isolates, or materials were specifically collected for this study, nor were any actions required from patients.

#### Sequence data

We sequenced 39 randomly selected strains of RT002 collected between 2013-2021. Total DNA was isolated from cultured bacteria for sequencing of these strains. In short, RT002 colonies were resuspended in Tris/EDTA (TE) buffer and heated at 100°C for 10 min according to the protocol (9). Chromosomal DNA was isolated using the QiaAmp blood and tissue kit (Qiagen) according to the instructions of the manufacturer. After preparation with the NebNext Ultra II DNA library prep kit for Illumina, DNA was sequenced at GenomeScan B.V. (Leiden, The Netherlands) on an Illumina NovaSeq6000. On average 3 million paired-end reads (read size 150 bp) were produced per sample, containing a minimum of 90% reads with a quality of 30 or more.

#### Ridom SeqSphere^+^ cgMLST and wgMLST

Ridom SeqSphere^+^ (version 6.0.2; Ridom GmbH, Münster, Germany) was run with default settings for quality trimming, *de novo* assembly, and allele calling. Quality trimming occurred at both 5’ and 3’ ends until reaching an average base quality of 30 with a length of 20 bases and a 120-fold coverage (10). *De novo* assembly was performed using the SKESA assembler version 2.3.0 (11) integrated in SeqSphere^+^ applying default settings. SeqSphere^+^ scanned for the defined genes using BLAST (12) with the criteria described previously (10).

Quality control of these assemblies was performed. The average assembly size was 4.2 Mb. The average N50 across the samples was 213448 bp (range: 18044– 501411 bp). The GC-content was consistent at 28.21% across samples. The average contig size was 106, with a range of 57 to 502 contigs. cgMLST analysis showed an average of 99.75% correctly assembled core loci. One sample formed an outlier with an average assembly size of 7.3Mb and a GC content of 30.6%, indicating possible contamination. However, the cgMLST analysis showed 99.7% correctly assembled core loci. Despite minor variations, the overall assembly quality was high and suitable for downstream analyses.

For further analysis, distance matrices, minimum spanning trees, and neighbor joining trees were produced using the integrated features within SeqSphere^+^ using the “pairwise ignoring missing values” option.

### Statistical analysis

Age was compared by a Wilcoxon rank-sum test and all other characteristics were compared by a Pearson’s chi-square test and in case of expected frequencies <5, a Fisher’s exact test was used. To compare the effect of RT002 and other ribotypes on clinical characteristics, separate multiple logistic regressions were performed on data restricted to only RT002 and the ribotype of interest, with age and RT as independent variables. A correction for multiple testing has not been performed, since we were focusing on general emerging patterns rather singling out individual significant results. A p value of <0.05 was therefore considered statistically significant. The statistical analysis was performed on IBM SPSS Statistics for Windows, Version: 28.0.1.0 (IBM Corp., Armonk, N.Y., USA).

### Data availability

All genome sequence data generated as part of this study were submitted to the NCBI/ENA under study number PRJEB81246.

## Results

### CDI in hospitalized patients due to RT002 is comparable with other non-hypervirulent ribotypes

In 2019, an increase in CDI with a clonal strain of RT002 was reported in Ireland. Prompted by this observation, we evaluated data collected by the Dutch national expertise center for *C. difficile* infections and noted a significant increase in CDI proportion due to RT002 from 5.4%-7.2% (between May 2009-April 2017) to 11.9% (between May 2017-April 2018) (Figure 1). The proportion of CDI due to RT002 remained between 9.6%-9.9% in the years following this period (May 2019-April 2021). We were wondering if this increase in proportion was accompanied with changes in clinical characteristics of patients. We therefore used the data provided by the Dutch national *C. difficile* surveillance program between May 2009 and April 2021, using 8080 PCR-ribotyped isolates from hospitalized patients in 24 hospitals in our analysis. Within this set, the RT002 group accounted for 611 isolates. The average proportion of RT002 was 7.5% making it the third most common RT behind RT014/020/295 (17.3%) and RT001 (10.3) in the Netherlands during that period.

**Figure 1:**
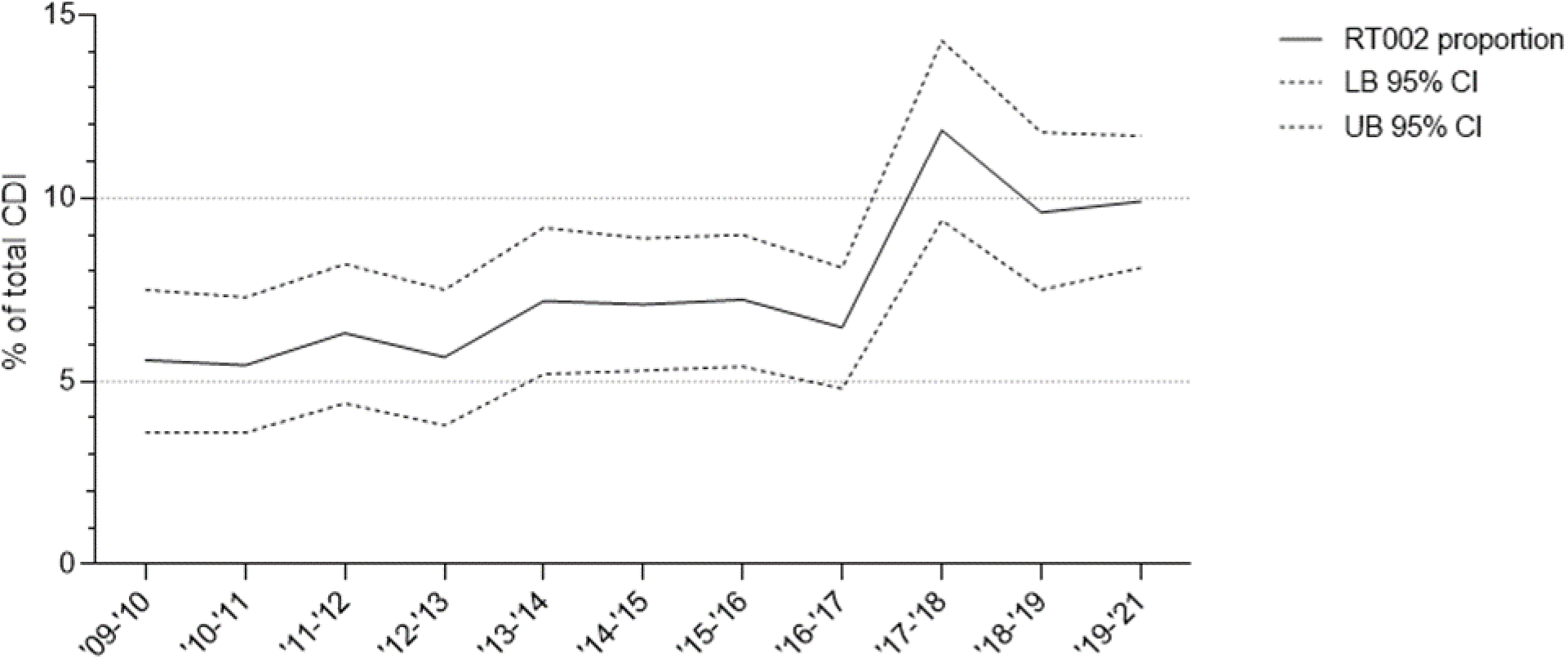
Proportion of RT002 of total CDI in time in the national sentinel surveillance samples. The lower boundary (LB) and upper boundary (UB) of the 95% confidence interval (CI) are shown with a dotted line.

We compared demographic data, clinical characteristics and 30-day outcome of patients with CDI caused by RT002 with five other ribotype groups (RT001, RT014/020/295, RT027, RT078/RT126 and all other RTs combined in the Others group) for the entire period (Table 1 and the detailed version in the supplementary file).There was no significant difference in age or gender between RT002 and the other groups, except for the RT001 group which had a higher average age. The RT002 group showed a higher severe CDI average (20.94%) than the RT001 group (16.98%, p=0.021), even after correcting for age (p=0.037). However, it showed a significantly lower severe CDI average when compared with the RT078/126 group (25.21%, p=0.041), even when corrected for age (p=0.034). RT002 group showed a lower complicated course average (14.13%) than the RT027 group (22.00%, p=0.045), but this was not significant after correction for age (p=0.054). It showed a higher complicated course average (14.13%, p=0.005, after correction for age p=0.009) and non-CDI mortality average (13.01%, p=0.004, after correction for age p=0.007) than the RT014/020/295 group (9.58% and 8.49%, respectively). The onset of symptoms of CDI in the RT002 group was more frequent at home compared with the RT027 group (31.82%, p= 0.003, after correction for age p=0.005) and the RT001 group (26.21% p <0.001, remains the same after correction). The antibiotic usage prior to CDI was significantly lower in the RT002 group (63.64%), compared with the RT001 group (76.81%, p<0.001, also after correction for age), the RT027 group (78.10% (p=0.004, after correction for age p 0.006) and the RT078/126 group (71.50%, p=0.002, also after correction for age). Overall, CDI due to RT002 is comparable to other non-hypervirulent ribotypes and occurs more frequently at home and is associated with less antibiotic usage.

**Table 1:**
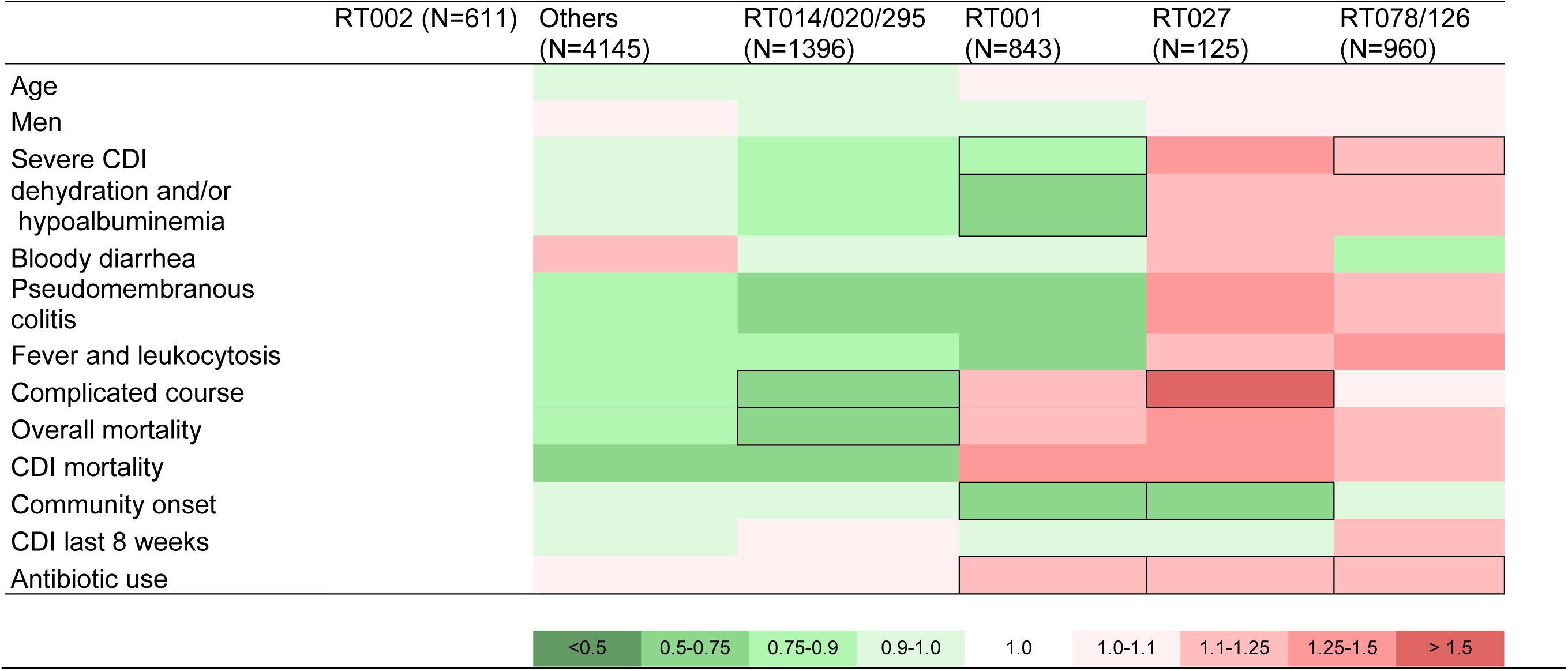
Comparison of clinical characteristics of patients with RT002 with other RTs (2009-2021). Heatmap plot demonstrating the pattern of clinical characteristics of RT002 compared with other RTs. When the incidence of a certain characteristics is lower compared to the incidence in RT002 the cell is given a certain shade of green When the incidence of a certain characteristics is higher compared to the incidence in RT002 the cell is given a certain shade of red. Significant differences are shown with bordered cells. The table with the exact values of these cells can be found in the supplementary files.

### CDI in hospitalized patients due to RT002 prior to the proportion increase is severe, comparable with RT027 and RT078/126 strains

Considering the proportion increase in 2017, we next split the dataset to assess whether clinical characteristics changed between these periods and created RT groups prior to the proportion increase (hereafter RT_09-17_; May 2009-April 2017) and RT groups after this increase in proportion (hereafter RT_17-21_; May 2017-April 2021) to see whether there were changes in demographics, clinical characteristics, 30-day outcome. The RT002_09-17_ group and the RT002_17-21_ group accounted for 348 isolates and 263 isolates, respectively. We observed a significant difference between RT002_09-17_ group (Table 2 and the detailed version in the supplementary file) in severe CDI average (27.24% vs 15.44%, p<0.001, also after correction for age) and dehydration and/or hypoalbuminemia average (16.72% vs 4.59%, p<0.001, also after correction for age) and lower incidence in males (44.80% vs 54.37%, p 0.018, after correction for age 0.016) compared to RT002_17-21_ group.

**Table 2:**
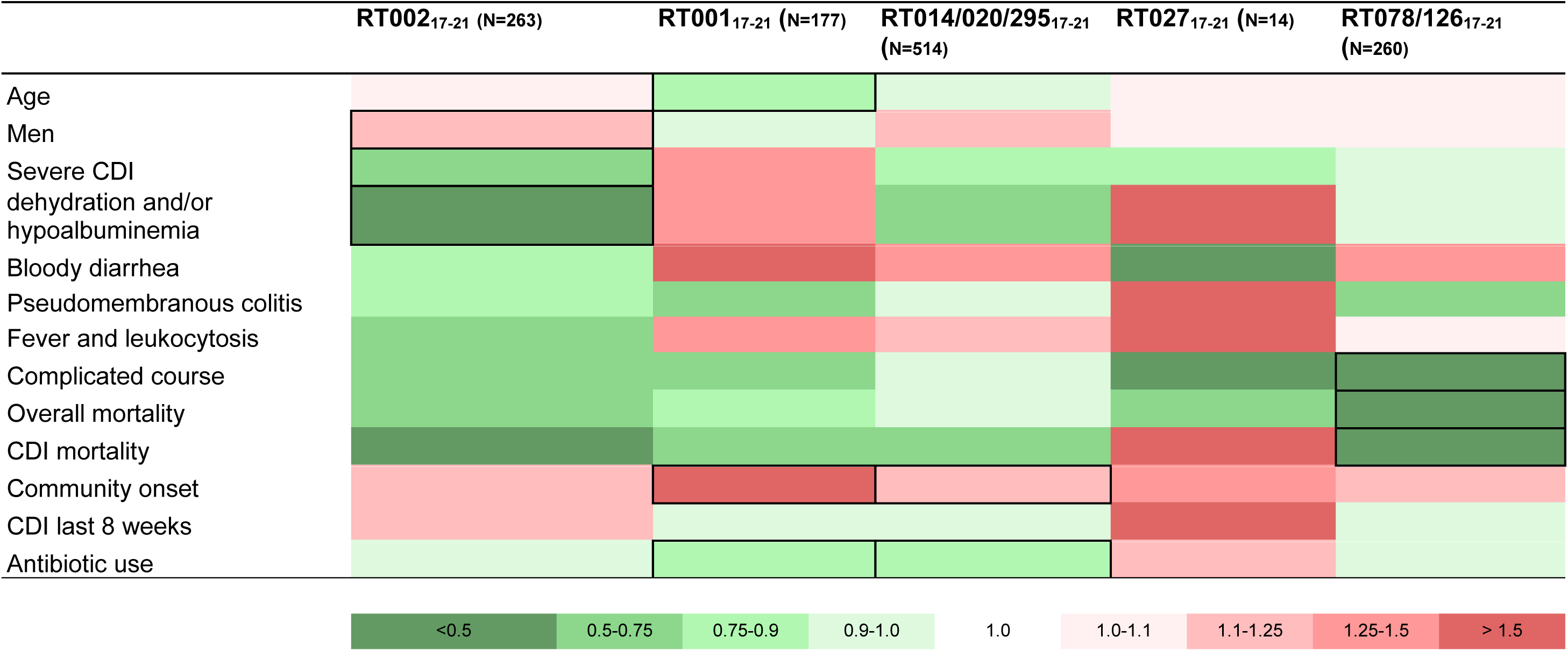
Comparison of clinical characteristics of patients between two groups of RT001, RT002, RT014/020/295, RT027 and RT078/126. Comparison of clinical characteristics of patients between two groups of RT001, RT002, RT014/020/295, RT027 and RT078/126. Heatmap plot demonstrating the pattern of clinical characteristics of RTs_09-17_ compared with RTs_17-21_. When the incidence of a certain characteristics is lower in RTs_17-21_ compared to the incidence in RTs_09-17_ the cell is given a certain shade of green. When the incidence of a certain characteristics is higher in RTs_17-21_ compared to the incidence in RTs_09-17_ the cell is given a certain shade of red. Significant differences are shown with bordered cells. The table with the exact values of these cells can be found in the supplementary files.

We noted that the characteristics of the RT002 isolates relative to the other RT groups appeared to differ between the periods May 2009-April 2017, and May 2017-April 2021. In particular, RT002 was more similar to HV-RTs prior to May 2017, whereas it was more similar to non-HV RTs over the period 2017-2021. The RT002_09-17_ group (Table S1A) showed a significantly higher rate of severe CDI, dehydration and/or hypoalbuminemia, fever and leukocytosis and complicated CDI course than non-HV RTs and the other ribotypes group. In contrast, the RT002_17-21_ group (Table S1B) had a comparable severe CDI average as the non-HV RTs and the other ribotypes group, but lower compared to HV-RTs (e.g. RT078/126_17-21_ group). (24.62%, p 0.006, also after correction for age). The average complicated CDI course of the RT002_17-21_ group was comparable with all ribotype groups.

When other ribotypes were analyzed in the same temporal subgroups, we did not observe such a trend (Table S2A-S4B). For instance, both RT001_09-17_ and RT014/020/295 _09-17_ appear to cause less severe disease than other ribotypes in the same period, whereas this was increased for RT002_09-17_.

### Dutch RT002 strains not related to the Irish outbreak strains, but certain Dutch strains are genetically related to each other

As the increase in proportion of CDI due to RT002 in the Netherlands appeared to precede the Irish reports, we investigated whether the Irish clonal strains were related to the Dutch RT002 strains. Thirty-nine randomly selected RT002 strains from the Netherlands between 2013-2021 were sequenced and compared to sequence data from 11 Irish strains (all collected in 2019), consisting of six Irish RT002 outbreak strains (from 2 known counties and 2 known labs and 1 unknown county and lab) and five randomly selected RT002 strains.

The Dutch RT002 strains showed no relatedness with the Irish outbreak strains in a cgMLST analysis using SeqSphere^+^ (Figure 2A). Five clusters were detected with cgMLST, one of which corresponded to the Irish clonal outbreak isolates that were included as controls in our analysis (MST Cluster 1). These outbreak strains (Irish-6 to Irish-11) formed a tight cluster with 0 allele difference in a cgMLST analysis. All Dutch RT002 isolates had >6 allele differences to these isolates. Two Dutch strains showed a 9-allele difference with cgMLST analysis. One Irish non-outbreak (Irish-5) strain was related to MST Cluster 1 and differed at 3 alleles. The other, non-outbreak Irish isolates (Irish-1 to Irish-4) were interspersed in the minimal spanning tree. The other 4 clusters consisted of 13 strains, comprising 10 strains from patients with mild CDI, 1 with severe CDI and 2 unknown. The strains from MST Cluster 2 differed 3-6 alleles (these include allele differences in the distance matrix, data not shown) from each other. MST Cluster 2 consisted of five strains, of which three were obtained from hospitalized patients from one hospital across different years (2013, 2017 and 2018). All three patients had the onset of their CDI outside the hospital: one patient in a long-term residential care facility and the other two at home. The other 2 strains were from 2 different hospitals in the same city at a distance of 50 km from the first three. One patient developed CDI at home, and the other in the hospital. All these five patients lived in a different city within a radius of 80 km from each other. All isolates were from patients with mild CDI with an uncomplicated CDI course. Cluster 3 was genetically related to MST Cluster 2, at a minimum distance of 8 alleles difference. The strains from MST Cluster 3 were from three different hospitals in different cities. The strains from MST Cluster 3 differed 1-5 alleles from each other. This group consisted of two mild CDI cases and 1 severe CDI case; all had an uncomplicated CDI course and developed CDI at home. All these patients lived in different cities within a radius of 130 km. MST Cluster 4 consisted of two Dutch strains from different cities and one Irish non-outbreak strain (Irish-4). The allele differences in these strains were 2-4 alleles. One patient had a mild CDI course and the CDI course of the others was not known. Lastly, MST Cluster 5 consisted of two strains from different cities at a distance of 270 km, with an allele difference of 5. These isolates were also from patients with a mild CDI course and developed CDI at home. In Figure 2B a neighbor joining tree is shown with the phylogenetic relationship of the Dutch strains with the Irish outbreak strains and where all clusters can be seen. Due to the close relatedness of MST Cluster 2 and MST Cluster 3, strain 2018-5 from MST Cluster 2 is shown separate from the other strains of MST Cluster 2.

**Figure 2.**
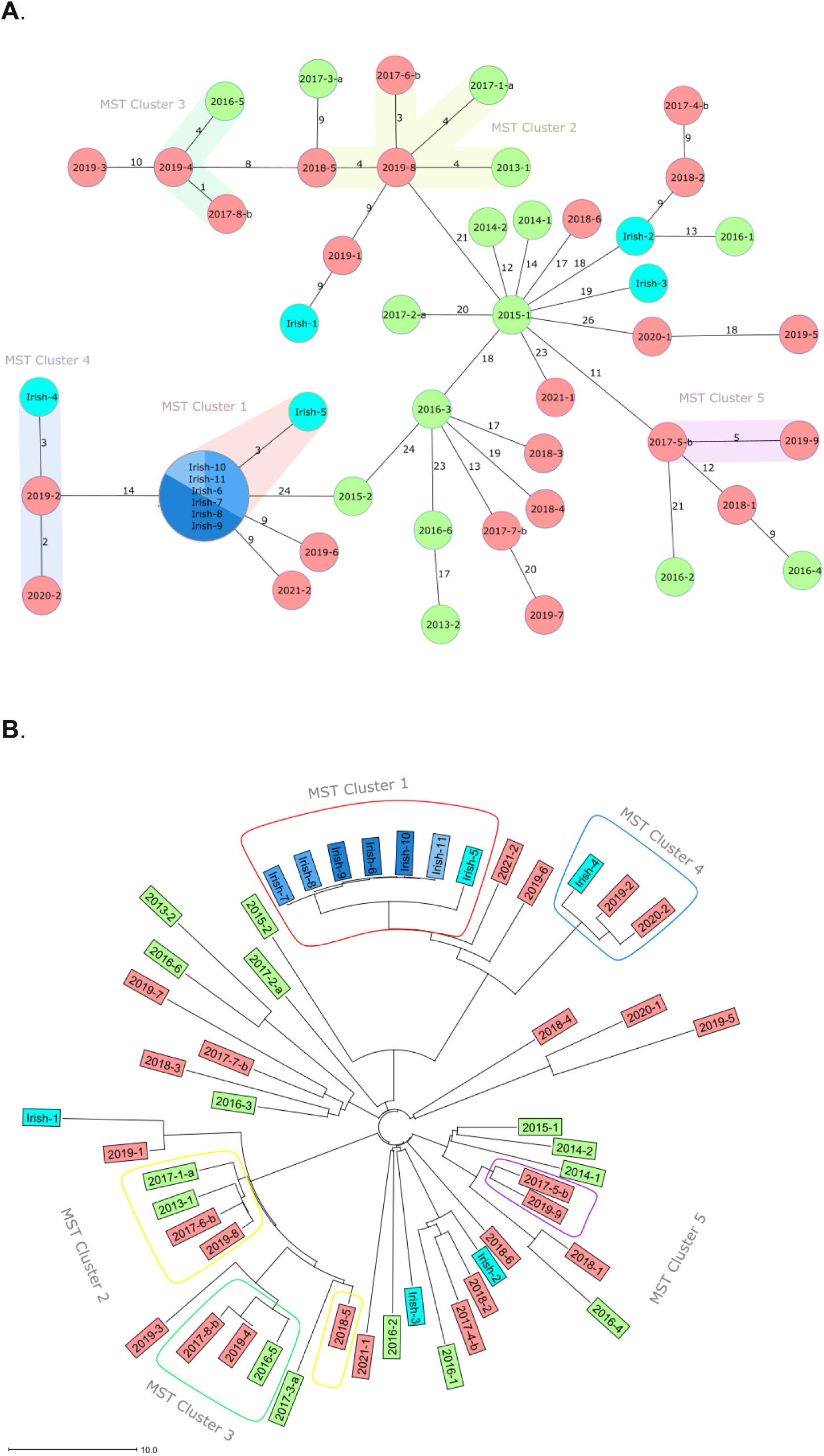
Analysis of relatedness of RT002 strains in this study. **A:** SeqSphere^+^ cgMLST analysis with minimum-spanning trees of Irish RT002 random strains (N=5, Irish 1-5, depicted as light blue) and clonal strains (N=6, Irish 6-11, depicted as several shades of blue inside of the biggest node) and random Dutch RT002 strains (N=39, RT002_09-17_ strains depicted as green and RT002 _17-21_ strains depicted as red). Dutch strains are stated as “year of isolation-sample.” Strains from 2017 end with “a” or “b” indicating strains prior or after the increase in proportion, respectively. One or more strains inside a node means that these strains have zero allele differences. The colored septations represent the laboratory from where the strains have been obtained. The numbers between each node correspond to the number of different alleles between the strains. The colored shadowing of circles represents a cluster with at most six allele differences that are genetically related. **B:** Neighbor joining tree based on SeqSphere^+^ cgMLST allele difference from 50 strains of RT002 also shown in Figure 2A. The distance is given as the absolute allelic difference. The colored encircling of strains represents a cluster with at most six allele differences that are genetically related. Cluster 1 (red shade), Cluster 2 (yellow shade), Cluster 3 (green shade), Cluster 4 (blue shade) and Cluster 5 (purple shade).

## Conclusion and Discussion

Our main goal was to study the increase in proportion of CDI due to RT002 in hospitalized patients to assess whether this was related to the Irish RT002 outbreak and whether this was accompanied by a change in clinical characteristics. We showed using cgMLST that the sequenced Dutch RT002 strains are not genetically related to the Irish outbreak strains. However, we found several clusters of RT002 strains in the Netherlands, suggesting clonal expansion. Though some of these clusters were recognized in the same hospital, most of these clusters comprised strains from different geographic locations at different time points, arguing against an outbreak or local transmission.

The overall CDI incidence was on average around 3.06 cases per 10.000 patient-days between 2010-2021, and CDI incidence due to RT002 was on average 0.22 cases per 10.000 patient-days. The proportion increase of RT002 that was noticed between May 2016-April 2017 and May 2017-April 2021 (Figure 1) preceded a general increased CDI incidence 1 year later (Figure S1).

The increase of CDI due to RT002 was not accompanied by an increase in severe disease. The clinical characteristics of hospitalized patients with CDI due to RT002 was similar to other non-hypervirulent ribotypes when assessed for the full 2009-2021 period. However, we observed that RT002 strains showed a higher CDI severity before the increased proportion, together with a higher complicated course and higher mortality. This was not observed in other ribotypes that were analyzed in the same temporal subgroups. In contrast, we have observed an opposite change in the RT001 group, where the RT001_17-21_ group showed more disease severity. If we look at the proportion of these two ribotypes in the Netherlands (Figure 1 based on data from the 14th Annual report of *C. difficile* 2019-2021 (13)), we also observe an inverse relation between disease severity and proportion rate. The proportion of RT001 was 26.5% between May 2009-May 2010 and decreased to 7% between May 2019-May 2021, whereas RT002 increased from 5.6% to 9.9% in the same period.

The reason for the increased proportion in combination with a decrease in disease severity in RT002 requires further research. Several causes for an increased proportion can be envisaged, such as mutations or acquisition of traits that confer advantages (such as antibiotic resistance, increased metabolic efficiency or enhanced survival in specific environments). A preliminary *in silico* analysis on the RT002 strains (data not shown) did not reveal alterations in or acquisition of genes associated with trehalose metabolism (14), or changes in the pathogenicity locus associated with increased virulence (15). Though a VanS-p.T349I mutation in RT027 has been associated with an increase in vancomycin MIC (16), antimicrobial susceptibility testing of RT002 strains carrying this mutation did not show elevated vancomycin MICs (<0.06 mg/L-0.06mg/L) (data not shown). In addition, no resistance determinants or phenotypic reduced susceptibility was found for metronidazole (data not shown). The higher rates of sporulation and germination of RT002 reported previously (4), (5) may possibly account for persistence of RT002 strains in the environment and could imply that its spores could germinate even in suboptimal environments. This could contribute to an increased proportion of RT002 CDI cases. However, these differences in proportion of CDI could be caused as well by other factors (e.g. antibiotic usage, dietary habits, change in infection prevention measurements and host factors).

We found clustering of strains belonging to *C. difficile* RT002 based on a cgMLST analysis using SeqSphere^+^, that uses ≤ 6 allele differences to define an MST Cluster. The average allele difference between RT002 strains from within and outside the Netherlands was 28.5 and 32.5 allele differences, respectively, in our previous study that also employed SeqSphere^+^ cgMLST (17). In the same study, strains from MLST Clade 2 had a lower intra-ribotype allele average (9 alleles) compared to MLST Clade 1 (114 alleles), to which RT002 belongs. Therefore, clonality should be easier to detect in comparison with a ribotype belonging to MLST Clade 2. For instance, we found a cluster (see Figure 2A) comprising 3 patients with community-onset CDI hospitalized with CDI due to RT002 in different years in the same hospital. It is possible that RT002 spores persist for several years in the hospital environment and occasionally infect patients who subsequently develop CDI after discharge. Another explanation could be a yet unknown common source outside the hospital. For instance, a study in 12 European countries showed that various PCR ribotypes, including RT002, were present on retail potatoes (18). The most likely explanation is the data from cgMLST for RT002 are limited for patient transmission studies.

During the preparation of this manuscript a study on the genetic characteristics of RT002/sequence type (ST) 8 was published (19). The authors observed close genetic relatedness among the studied ST8 genomes from different European countries, acquisition of a varied array of antimicrobial resistance and presence of numerous mobile elements. Furthermore, they found clonal isolates across distinct One Health sectors, geographic location and time periods. These results suggest that isolates of RT002 are genetically more closely related than other RT lineages and that related isolates can be detected independently from the geographical origin. Clonality (e.g. in terms of cgSNP differences) does not necessarily indicate a direct transmission.

The strengths of this study are the 12 years of available data from hospitals in different geographic regions that participated in the sentinel CDI surveillance and the high sample size. There are also a few limitations, such as the small numbers of sequenced strains and uncommon ribotype groups with a low incidence after splitting the data set in two groups (e.g. PCR ribotype 027), which limits statistical power. Therefore, we only corrected for age as a confounder. Furthermore, only the location of CDI symptoms onset, and not the location of *C. difficile* acquisition, were recorded. The national CDI surveillance does not collect data about comorbidities that may affect the clinical outcome. However, CDI attributable mortality is included in the surveillance and data analysis. Furthermore, retrospective interpretation on possible epidemiologically related clusters can be problematic (e.g. 3 strains from MST Cluster 2 from the same hospital).

This paper highlights that the results of a cgMLST should be interpreted with care with respect to understanding pathways of patient transmission. Without the context of wider sequencing data, epidemiological data and metadata, cgMLST cannot differentiate between links attributed to widely disseminated clonal strains (for example in food or water) and those related to site specific chains of transmission of infection (local outbreaks) (17). This differentiation is essential for appropriately targeted interventions. The usage of cgMLST in combination with a national sentinel surveillance system is important to detect the emergence of certain strains over the years. It is, therefore, of great importance to continue epidemiological surveillance and genetic characterization of C. *difficile*.

## Data Availability

All data produced in the present study are available upon reasonable request to the authors, or are available from public repositories through the information provided in this manuscript.

## Notes

### Competing Interest Statement

The authors have declared no competing interest.

### Funding Statement

This study did not receive any funding.

### Author Declarations

According to the Dutch Medical Research Involving Human Subjects Act (WMO), the National Surveillance Progamme of CDI was considered exempt from review by an Institutional Review Board.

