## Supplementary material for "Increase of *Clostridioides difficile* PCR ribotype 002 infection coincides with decreased disease severity in hospitalized patients in the Netherlands": Baktash_RT002 Manuscript_SupplementaryInformation.pdf

### SUPPLEMENTARY INFORMATION

Figure S1: CDI cases per 10.000 patient-days shown for all ribotypes (depicted in blue) and for RT002 (depicted in red).

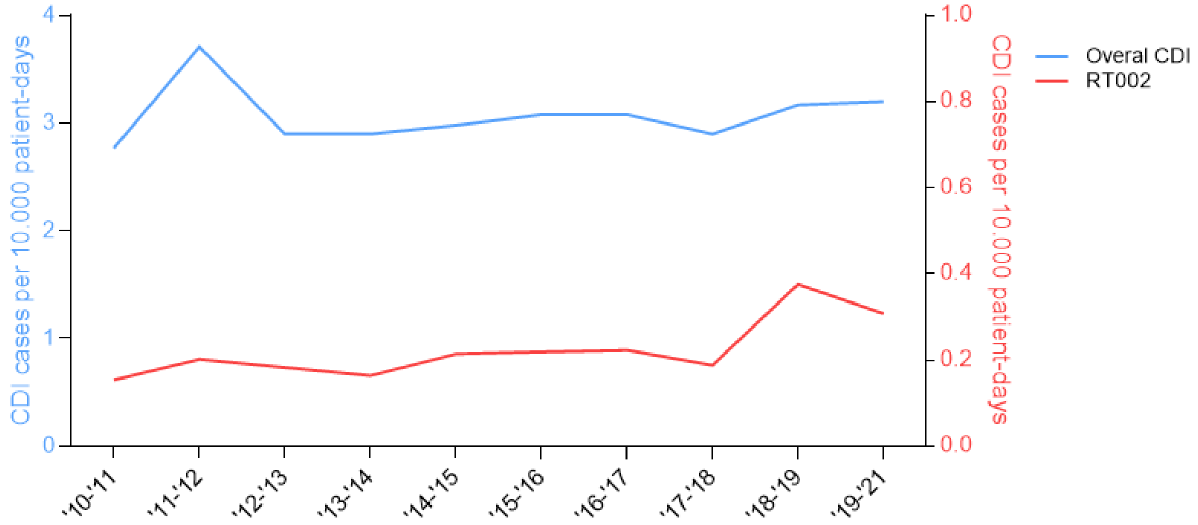

**Table S1A:** Comparison of clinical characteristics of patients with RT002$_{09-17}$ with other RTs$_{09-17}$

| | RT002$_{09-17}$ (N=348) | Others$_{09-17}$ (N=2739) | RT014/020/295$_{09-17}$ (N=882) | RT001$_{09-17}$ (N=666) | RT027$_{09-17}$ (N=111) | RT078/126$_{09-17}$ (N=700) |
|---|---|---|---|---|---|---|
| Age | | | | | | |
| Men | | | | | | |
| Severe CDI | | | | | | |
| dehydration and/or hypoalbuminemia | | | | | | |
| Bloody diarrhea | | | | | | |
| Pseudomembranous colitis | | | | | | |
| Fever and leukocytosis | | | | | | |
| Complicated course | | | | | | |
| Overall mortality | | | | | | |
| CDI mortality | | | | | | |
| Community onset | | | | | | |
| CDI last 8 weeks | | | | | | |
| Antibiotic use | | | | | | |

| <0.5 | 0.5-0.75 | 0.75-0.9 | 0.9-1.0 | 1.0 | 1.0-1.1 | 1.1-1.25 | 1.25-1.5 | > 1.5 |
|---|---|---|---|---|---|---|---|---|

Heatmap plot demonstrating the pattern of clinical characteristics of RT002$_{09-17}$ compared with other RTs$_{09-17}$. When the incidence of a certain characteristics is lower in RTs$_{09-17}$ compared to the incidence in RT002$_{09-17}$ the cell is given a certain shade of green. When the incidence of a certain characteristic is higher in RTs$_{09-17}$ compared to the incidence in RT002$_{09-17}$ the cell is given a certain shade of red. Significant differences are shown with bordered cells. The table with the exact values of these cells can be found in the supplementary files.

**Table S1B:** Comparison of clinical characteristics of patients with $RT002_{17\text{-}21}$ with other $RTs_{17\text{-}21}$

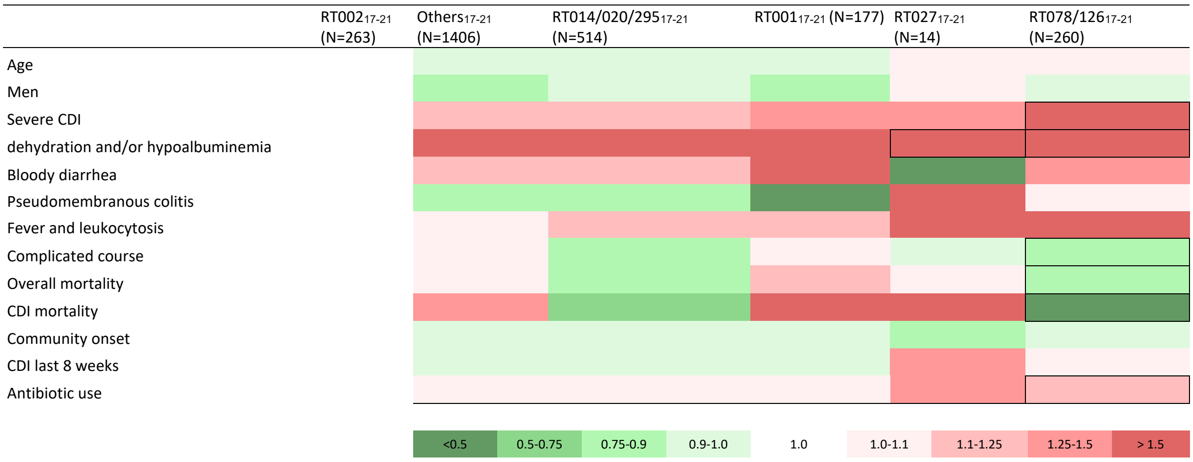


Heatmap plot demonstrating the pattern of clinical characteristics of $RT002_{17\text{-}21}$ compared with other $RTs_{17\text{-}21}$. When the incidence of a certain characteristics is lower in $RTs_{17\text{-}21}$ compared to the incidence in $RT002_{17\text{-}21}$ the cell is given a certain shade of green. When the incidence of a certain characteristic is higher in $RTs_{17\text{-}21}$ compared to the incidence in $RT002_{17\text{-}21}$ the cell is given a certain shade of red. Significant differences are shown with bordered cells. The table with the exact values of these cells can be found in the supplementary files.

**Table S2A:** Comparison of clinical characteristics of patients with RT001$_{09-17}$ with other RTs$_{09-17}$

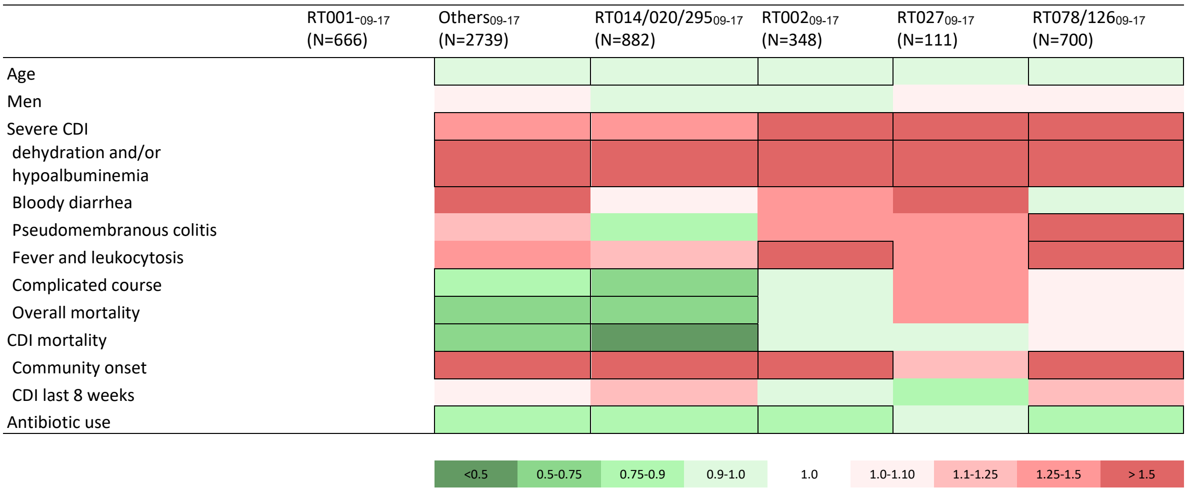


Heatmap plot demonstrating the pattern of clinical characteristics of RT001$_{09-17}$ compared with other RTs$_{09-17}$). When the incidence of a certain characteristics is lower in RTs$_{09-17}$ compared to the incidence in RT001$_{09-17}$ the cell is given a certain shade of green. When the incidence of a certain characteristic is higher in RTs$_{09-17}$ compared to the incidence in RT001$_{09-17}$ the cell is given a certain shade of red. Significant differences are shown with bordered cells. The table with the exact values of these cells can be found in the supplementary files.

**Table S2B:** Comparison of clinical characteristics of patients with RT001$_{17\text{-}21}$ with other RTs$_{17\text{-}21}$

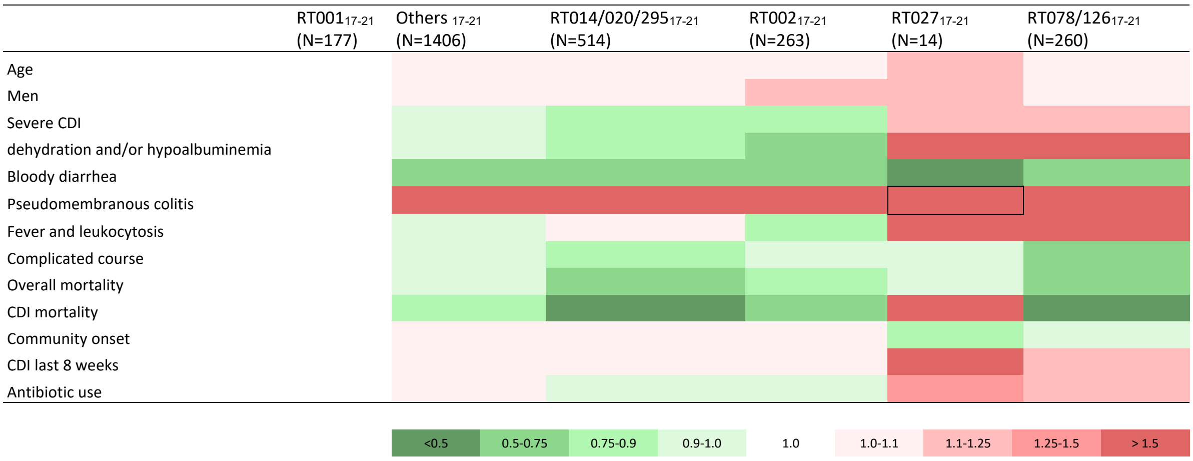


Heatmap plot demonstrating the pattern of clinical characteristics of RT001$_{17\text{-}21}$ compared with other RTs$_{17\text{-}21}$. When the incidence of a certain characteristics is lower in RTs$_{17\text{-}21}$ compared to the incidence in RT001$_{17\text{-}21}$ the cell is given a certain shade of green. When the incidence of a certain characteristic is higher compared to the incidence in RT001$_{17\text{-}21}$ the cell is given a certain shade of red. Significant differences are shown with bordered cells. The table with the exact values of these cells can be found in the supplementary files.

**Table S3A:** Comparison of clinical characteristics of patients with RT014/020/295$_{09\text{-}17}$ to other RTs$_{09\text{-}17}$

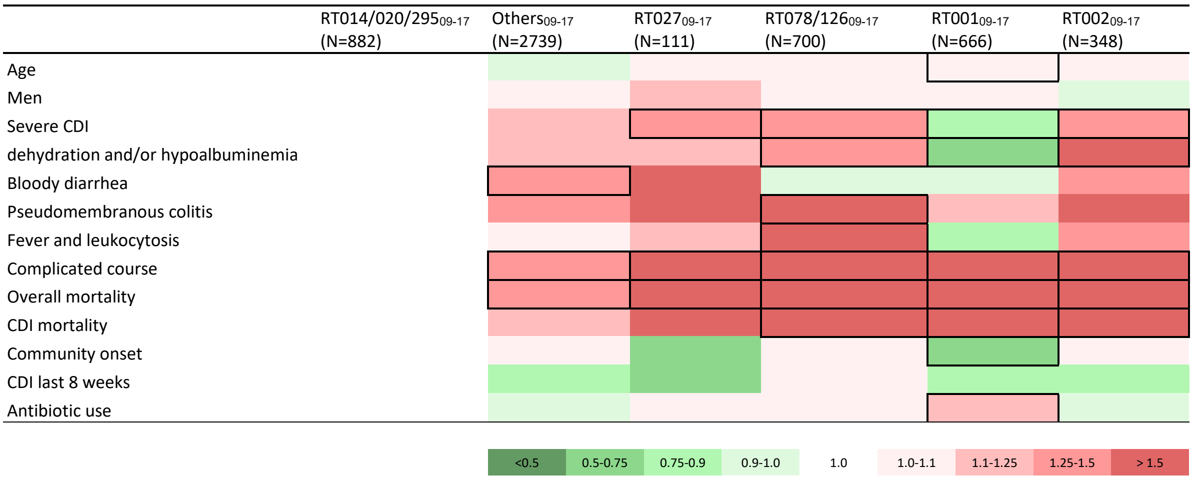


Heatmap plot demonstrating the pattern of clinical characteristics of RT014/020/295$_{09\text{-}17}$ compared with other RTs$_{09\text{-}17}$). When the incidence of a certain characteristics is lower in RTs$_{09\text{-}17}$ compared to the incidence in RT014/020/295$_{09\text{-}17}$ the cell is given a certain shade of green. When the incidence of a certain characteristic is higher in RTs$_{09\text{-}17}$ compared to the incidence in RT014/020/295$_{09\text{-}17}$ the cell is given a certain shade of red. Significant differences are shown with bordered cells. The table with the exact values of these cells can be found in the supplementary files.

**Table S3B:** Comparison of clinical characteristics of patients with RT014/020/295$_{17-21}$ with other RTs$_{17-21}$

| | RT014/020/295$_{17-21}$ (N=514) | Others$_{17-21}$ (N=1406) | RT027$_{17-21}$ (N=14) | RT078/126$_{17-21}$ (N=260) | RT001$_{17-21}$ (N=177) | RT002$_{17-21}$ (N=263) |
|---|---|---|---|---|---|---|
| Age | | | | | | |
| Men | | | | | | |
| Severe CDI | | | | | | |
| dehydration and/or hypoalbuminemia | | | | | | |
| Bloody diarrhea | | | | | | |
| Pseudomembranous colitis | | | | | | |
| Fever and leukocytosis | | | | | | |
| Complicated course | | | | | | |
| Overall mortality | | | | | | |
| CDI mortality | | | | | | |
| Community onset | | | | | | |
| CDI last 8 weeks | | | | | | |
| Antibiotic use | | | | | | |

| | | | | | | | | |
|---|---|---|---|---|---|---|---|---|
| <0.5 | 0.5-0.75 | 0.75-0.9 | 0.9-1.0 | 1.0 | 1.0-1.1 | 1.1-1.25 | 1.25-1.5 | > 1.5 |

Heatmap plot demonstrating the pattern of clinical characteristics of RT014/020/295$_{17-21}$ compared with other RTs$_{17-21}$. When the incidence of a certain characteristics is lower in RTs$_{17-21}$ compared to the incidence in RT014/020/295$_{17-21}$ the cell is given a certain shade of green. When the incidence of a certain characteristic is higher compared to the incidence in RT014/020/295$_{17-21}$ the cell is given a certain shade of red. Significant differences are shown with bordered cells. The table with the exact values of these cells can be found in the supplementary files.

**Table S4A:** Comparison of clinical characteristics of patients with RT078/126$_{09\text{-}17}$ to other RTs$_{09\text{-}17}$

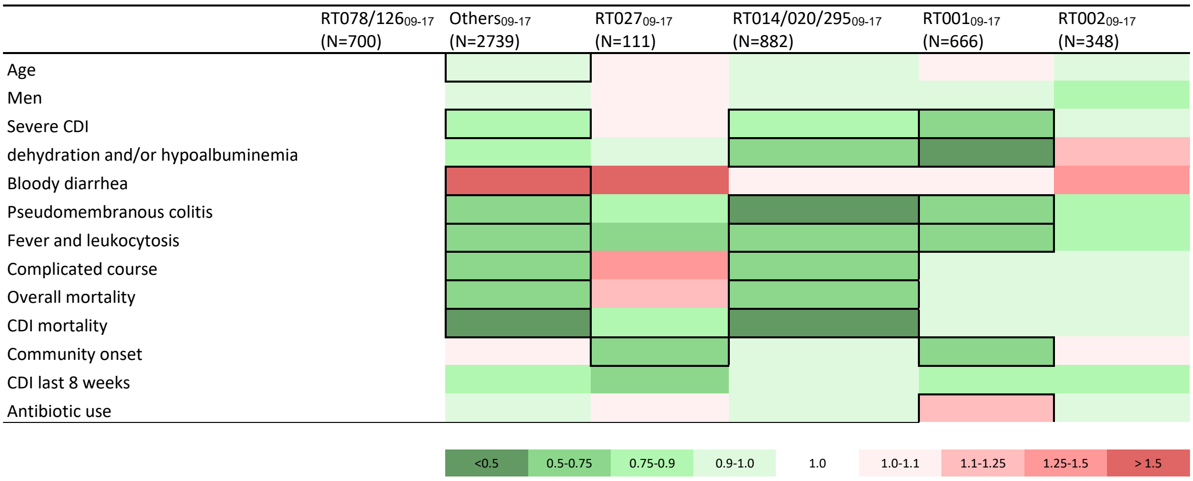


Heatmap plot demonstrating the pattern of clinical characteristics of RT078/126$_{09\text{-}17}$ compared with other RTs$_{09\text{-}17}$). When the incidence of a certain characteristics is lower in RTs$_{09\text{-}17}$ compared to the incidence in RT078/126$_{09\text{-}17}$ the cell is given a certain shade of green. When the incidence of a certain characteristic is higher in RTs$_{09\text{-}17}$ compared to the incidence in RT078/126$_{09\text{-}17}$ the cell is given a certain shade of red. Significant differences are shown with bordered cells. The table with the exact values of these cells can be found in the supplementary files.

**Table S4B:** Comparison of clinical characteristics of patients with RT078/126$_{17\text{-}21}$ with other RTs$_{17\text{-}21}$

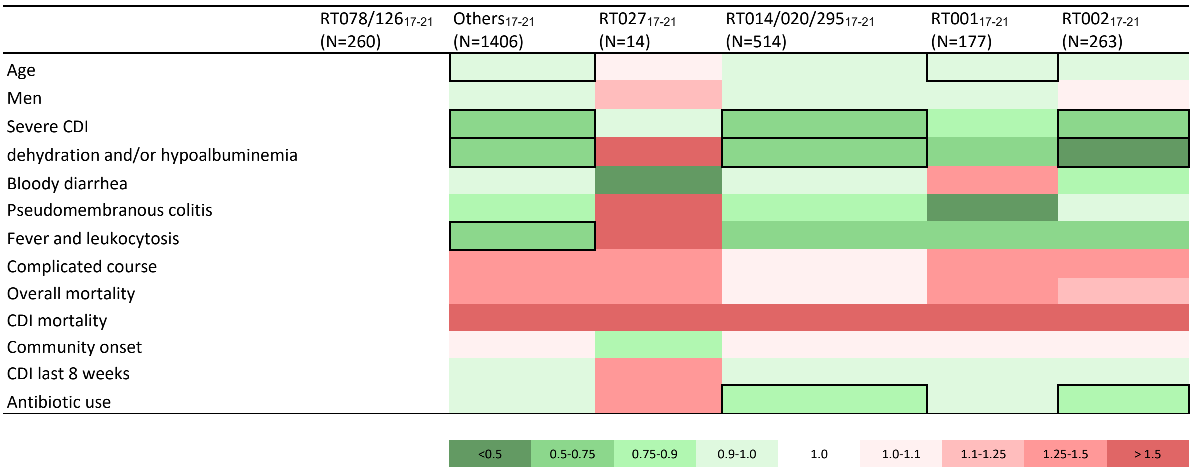


Heatmap plot demonstrating the pattern of clinical characteristics of RT078/126$_{17\text{-}21}$ compared with other RTs$_{17\text{-}21}$. When the incidence of a certain characteristics is lower in RTs$_{17\text{-}21}$ compared to the incidence in RT078/126$_{17\text{-}21}$ the cell is given a certain shade of green. When the incidence of a certain characteristic is higher compared to the incidence in RT078/126$_{17\text{-}21}$ the cell is given a certain shade of red. Significant differences are shown with bordered cells. The table with the exact values of these cells can be found in the supplementary files.
